# Multiplex PCR Assays for Identifying All Major SARS-CoV-2 Variants

**DOI:** 10.1101/2021.10.28.21263107

**Authors:** Ryan J. Dikdan, Salvatore AE Marras, Amanda P. Field, Alicia Brownlee, Alexander Cironi, D. Ashley Hill, Sanjay Tyagi

## Abstract

Variants of Concern (VOC) of SARS-CoV-2, including Alpha, Beta, Gamma, Delta, and Omicron threaten to prolong the pandemic leading to more global morbidity and mortality. Genome sequencing is the mainstay of tracking the development and evolution of the virus, but is costly, slow, and not easily accessible. A multiplex qRT-PCR assay for SARS-CoV-2 was developed, which identifies all VOC as well as other mutations of interest in the viral genome, eight mutations total, using single nucleotide discriminating molecular beacons in a two-tube assay. The presented variant molecular beacon assay showed a limit of detection of five copies of the viral RNA, with 100% specificity. Twenty-six SARS-CoV-2 positive patient samples were blinded and tested using this assay. When testing patient samples, the assay was in full agreement with results from deep sequencing with a sensitivity and specificity of 100% (26/26). We have used our design methodology to rapidly design an assay which detects the new Omicron variant. This Omicron assay was used to accurately identify this variant in 17 of 33 additional patient samples. These qRT-PCR assays identify all currently circulating VOC of SARS-CoV-2 as well as other important mutations in its Spike protein coding sequence. These assays can be easily implemented on broadly available five-color thermal cyclers and will help track the spread of these variants.

## Introduction

The COVID-19 pandemic has changed the world, leading to at least 5.3 million deaths and many permanent injuries, with over 271 million cases worldwide as of 20 December 2021 (https://www.who.int/emergencies/diseases/novel-coronavirus-2019, date of last access: 12/20/2021). Despite advances in diagnosis and vaccination, the emergence of SARS-CoV-2 variants threatens to keep the pandemic going^1^. The World Health Organization and the US Centers for Disease Control (CDC) (https://www.cdc.gov/coronavirus/2019-ncov/variants/variant-info.html, date of last access: 12/20/2021) have identified SARS-CoV-2 Variants of Concern (VOCs), which lead to increased disease severity (https://www.gov.uk/government/publications/nervtag-paper-on-covid-19-variant-of-concern-b117, date of last access: 12/20/2021), increased transmission^2^, and immune/vaccine evasion^3–5^. Variants of Interest (VOIs) have also been identified that present theoretical risks because they possess mutations similar to the mutations in the VOC. Specific frequently occurring mutations have also been identified which can affect therapeutic antibody treatments for patients infected with such variants^6^. These new variants and substitutions necessitate new tools for their detection and tracking^7^.

Currently, the most common technique to identify, classify, and track variants of SARS-CoV-2 is deep sequencing^8,9^. While sequencing is accurate and can identify each mutation present in a sample, it is costly, slow, and requires specialized instruments and interpretation when compared to other genotyping techniques, such as Polymerase Chain Reaction (PCR)^1^. Although, PCR assays target preselected mutations, they are cheaper and more accessible than sequencing for the genotyping of SARS-CoV-2, and they yield faster results. These methods may permit better tracking of variants as they spread throughout populations over time, especially in resource limited settings^1^.

Several SARS-CoV-2 variant genotyping PCR assays have been developed. They utilize TaqMan probes as those from Thermo Fisher Scientific (Waltham, MA), sloppy molecular beacon probes followed by melting temperature analysis^9^, and several other strategies such as PerkinElmer’s PKamp (Waltham, MA), and Aldatu Biosciences (Watertown,MA). However, most of these assays either target single mutations per reaction or lack the appropriate number of targets to thoroughly characterize SARS-CoV-2. Simple assays that detect multiple mutations at the same time are needed for identification of circulating VOCs, VOIs, and for the identification of specific mutations.

Our approach is to create a multiplex assay for SARS-CoV-2 variants by exploiting the superior selectivity and self-quenching characteristics of molecular beacons. Molecular beacons can be designed to selectively bind to a mutant target sequence, while avoiding the wild-type sequence, which differs from the mutant by a single nucleotide substitution. The interaction of a molecular beacon with its target is inherently more specific than linear oligonucleotides that do not have a stem and loop structure, because target binding requires the dissociation of the stem, which is thermodynamically costly^10^. The utility of this discriminatory power has been previously demonstrated in real-time PCR for a number of applications^11,12^.

Here we present a two-tube multiplex qRT-PCR assay that identifies current VOCs by detecting eight different mutations in the SARS-CoV-2 spike protein. We selected mutations that have been shown to increase immune escape, avoid neutralization, and increase transmissibility. Targeting these causative mutations is fruitful, because these strains may mutate and lose certain coincidental mutations, but mutations that increase transmission and immune evasion are likely to be maintained. Also, by targeting these types of mutations, it is possible to detect new variants that arise from combinations of previously confirmed mutations, as is the case of VOC Omicron. We also describe a molecular beacon design process that can be used to rapidly produce allele discriminating assays for new variants as they arise. As a proof of this design process, we created a new assay within two weeks for accurately identifying the Omicron variant.

## Materials and Methods

### Synthetic RNA targets

RNA standards of the SARS-CoV-2 whole genome and variant spike proteins were provided by Exact Diagnostics through Bio-Rad Laboratories, Hercules, CA (catalog numbers COV019, COV019CE, COVA, COVB, COVE, COVG). Additional spike protein RNA sequences, such as those containing the E484Q and T478K mutations and the omicron spike RNA, were generated from a double-stranded synthetic DNA (gBlock) from Integrated DNA Technologies (Coralville, IA). A 5’-T7 promoter was appended to it, which enabled transcription by T7 RNA polymerase, which was performed using the HiScribe™ T7 High Yield RNA Synthesis Kit (New England Biolabs, Ipswich, MA). The RNA produced was purified with a Monarch RNA Cleanup Kit, 50 µg, (New England Biolabs) and by denaturing polyacrylamide gel electrophoresis, eluting the band of correct size in gel elution buffer (400 mM NaCl, 20 mM Tris-HCl (pH 8.0), and 1 mM EDTA). The concentration of the eluted RNAs was determined by a NanoDrop spectrophotometer (Thermo Fisher Scientific).

### Primer Design

Primers were designed by using the primer3 server^13^. 200 nucleotides surrounding the mutation of interest on either side of the SARS-CoV-2 genome was used as input, the target-binding region of the molecular beacon was used as the hybridization probe, and the amplicon size was set between 50 to 300 nucleotides. The sequences of all the primers that were used in the assay are shown in Table 1.

**Table 1.**
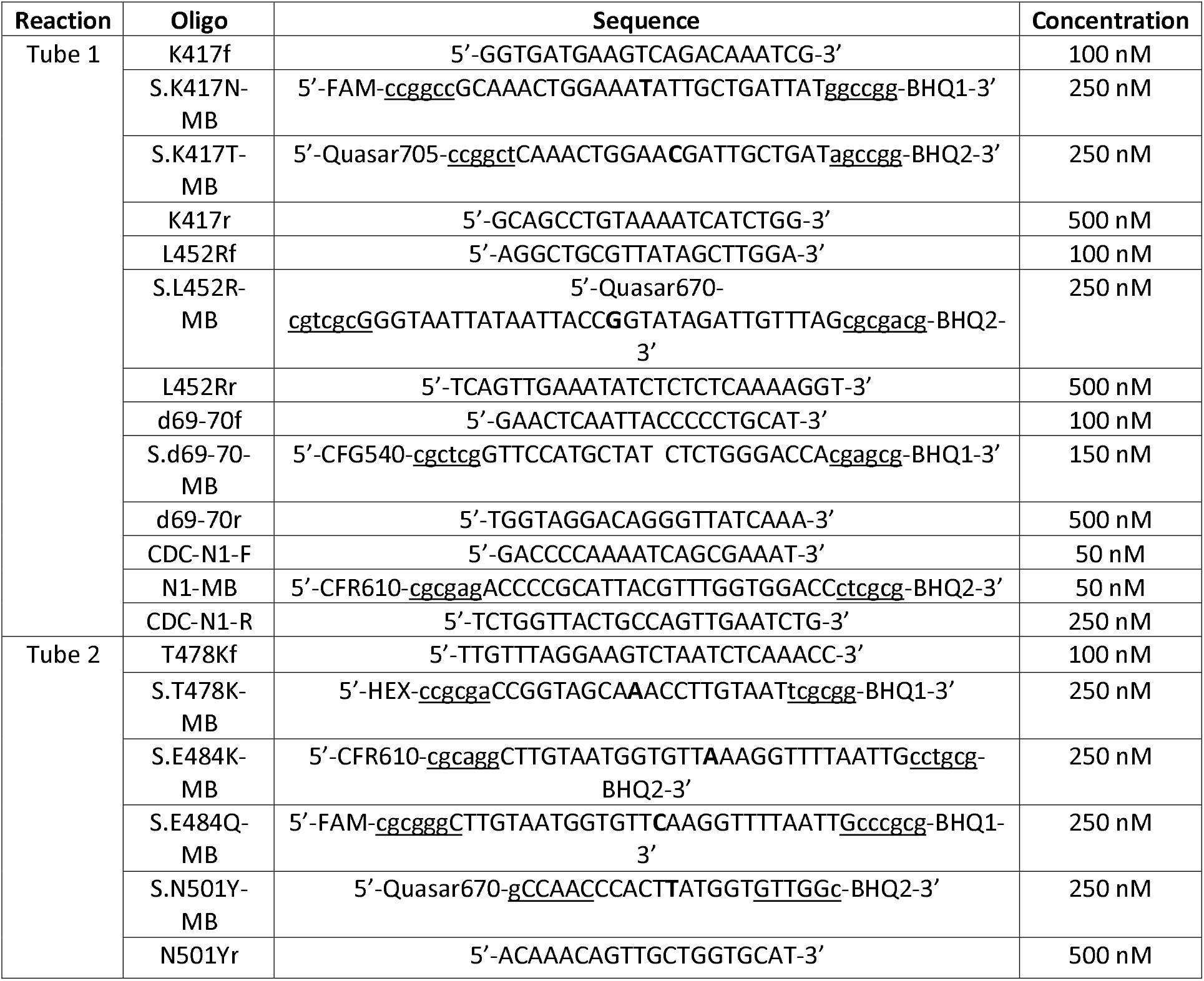
Primers and probes used in multiplex assay. Sequences are shown in the 5’->3’ direction. Upper case text indicates that it is a binding sequence, underlined text forms the hairpins of the molecular beacons, and bold letters indicate the point mutation nucleotides being identified by that molecular beacon. A gap indicates where the deletion is for the d69-70 molecular beacon. Abbreviations used: CFG = Cal Fluor Gold 540, CFR = Cal Fluor Red 610, BHQ = Black Hole Quencher

### Design and characterization of single nucleotide discriminating molecular beacons

A key consideration in the design of single-nucleotide discriminating molecular beacons is that the melting temperature (T_m_) of the probe-target hybrid with the perfect target (in the present case, the mutant sequence) is minimally above, only 3 to 7 °C, the annealing temperature of the PCR (which is the temperature at which fluorescence will be monitored). This makes it so that the mismatched hybrid (in the present case, the wild-type sequence) will not appreciably bind to the molecular beacon at the fluorescence monitoring temperature. Furthermore, the melting temperature of the stem of the molecular beacon should be above the detection temperature.

We designed our primers so that they will function at the PCR annealing temperature of 58 °C. We selected a region of sequence around the site of each mutation such that the T_m_ of the perfect probe-target hybrid was above the annealing temperature, but that the mismatched single-nucleotide polymorphism had a T_m_ lower than the annealing temperature. These T_m_‘s were predicted by using the DINAMelt^14^ two-state melting hybridization server with the following parameters: 0.25 µM oligonucleotide concentration, [Na^+^] = 60 mM, [Mg^2+^] = 3 mM, for DNA at 58 °C.

After deciding upon the target binding region, a hairpin stem was created by appending 5 or 6 nucleotides on either side (with 4 to 5 G:C pairs) such that they are complementary to each other, but not complementary to the target. In addition, we ensured that the 5’ nucleotide is not a guanosine, because that could lead to fluorescence quenching due to the 5’ position of the fluorophore. These sequences were then ordered from either LGC Biosearch Technologies (Petaluma, CA) or from Eurofins (Luxembourg City, Luxembourg). They were purified by HPLC through a PRP-3 Reversed Phase HPLC Column (Hamilton Company, Reno, NV) in a Beckman System Gold HPLC System (Fullerton, CA). The desired fractions were then ethanol precipitated and resuspended in water, and their concentration was determined by NanoDrop spectrophotometer. The sequences of all molecular beacons that were used in the assay are shown in Table 1.

To determine if the designed sequences could sufficiently discriminate the alleles at the annealing temperature of the PCR assay, we measured the fluorescence of the molecular beacons in the presence of either a mutant target, a wild-type target or, no target, under the buffer conditions that are expected to be present during PCR. Each 25 µL thermal denaturation reaction contained 20 mM Tris-HCl (pH 8.4), 50 mM KCl, 1.5 mM MgCl_2,_ 1 µM oligonucleotide target, and 0.4 µM molecular beacon. The fluorescence intensity of each reaction was measured at every degree in the appropriate fluorescence channel as the temperature was lowered from 95 °C to 30 °C (decreasing 1 °C/10 seconds) in a Bio-Rad CFX96 Touch Real-Time PCR Detection System.

Melt curve signals were normalized, for each molecular beacon. The difference between a normalization point and the other melt curve fluorescent signals was subtracted from the raw fluorescent intensity values and then all values in that channel were divided by the maximum intensity for that molecular beacon, to derive relative fluorescence units, which range between 0 and 1. These data are shown in Figure S1.

### Optimizing the concentrations of the qRT-PCR multiplex primers and molecular beacons

qRT-PCR was performed with TaqMan Fast Virus 1-Step Master Mix (No ROX) to optimize conditions, and TaqPath™ 1-Step Multiplex Master Mix (No ROX) (both from Applied Biosystems, Waltham, MA) for final experiments and patient samples.

Asymmetric PCR was performed for each amplicon, where the primer generating the strand complementary to the molecular beacon (labeled reverse) was in 5-fold excess over the other primer (labeled forward). All molecular beacons were designed in the same forward strand direction. G:T mismatches can be discriminated from each other by switching the strand being interrogated to the complementary strand^15^. The molecular beacon and primer concentrations were adjusted by trial and error, for sufficient amplification and signal. The final working concentrations of each of the components for each tube are listed in Table 1 along with their sequences.

Each reaction had a final volume of 20 µL comprised of TaqPath or TaqMan Master 1-Step Mix (No ROX), water, and primers and probes as previously described. 15 µL of this complete master mix was pipetted into each PCR tube, and then 5 µL of sample was added and mixed, before running on the thermal cycler.

### Assay sensitivity and specificity

The sensitivity of the assay was determined by preparing dilution series of *in vitro* transcribed targets, which were diluted 10-fold from 1,000 copies/µL to 0.1 copies/µL. 5 µL of each of these dilutions were combined with 15 µL of completed master mix per reaction. The tubes were sealed, briefly shaken, spun down at 800 g for 1 min. and then run on a Bio-Rad CFX96 Touch Real-Time PCR Detection System with the TaqPath or TaqMan Fast specified thermal cycler conditions, and with an annealing temperature of 58 °C. Specificity of the assay was determined by comparing the known sequences of the *in vitro* transcribed targets with the signals in these dilution series. For patient samples, sensitivity and specificity were determined by comparing this assay’s results to sequencing results, which are the gold standard.

### Collection of patient samples and sequencing

Patient mid-nasal swabs were collected for clinical testing in 1 ml of DNA/RNA Shield Reagent (Zymo, Irvine, CA). 400 µL aliquots were used for RNA extraction using the Kingfisher Flex magnetic bead extraction system (Thermo Fisher Scientific) and the Zymo Quick RNA/DNA Magbead kit (Zymo) per manufacturer’s instructions. qRT-PCR screening was performed using 2 µL of purified RNA, TaqPath 1-step Multiplex Master Mix (No ROX) and an in-house validated multiplex screening assay. This screening assay, outlined in Figure S2, detects conserved sequences in the *N* and *RdRp* genes of SARS-CoV-2, along with human *ACTB (β-actin)* mRNA, with high sensitivity. The primers and molecular beacons targeting regions in the *RdRp, N*, and human *ACTB* genes were modified and adapted from previously validated assays^16^ (https://www.cdc.gov/coronavirus/2019-ncov/lab/rt-pcr-panel-primer-probes.html, date of last access: 12/7/2021), and are listed in Table 2. Samples were run on a Quantstudio 7 (Thermo Fisher Scientific). Amplicon sequencing was performed using the Ion AmpliSeq SARS-CoV-2 Insight Research Panel (Ion Torrent, Guilford, CT). Each library contained six samples, one positive control sample and one no template control. Barcoded libraries were prepared using the Ampliseq Kit for Chef DL8 and Ion 530 Kit-Chef (Ion Torrent). Sixteen libraries were combined to run on a single Ion 530 chip. Templates were prepared using the Ion 530 Chef Reagents template kit. Sequencing was performed on the Ion S5XL instrument with Ion S5 Sequencing Solutions and Reagents (Ion Torrent). Sequence data was analyzed for on-target and mean depth of coverage using the SARS_COV_2_Coverage Analysis plug-in v1.3.0.2 (Ion Torrent). FASTA files were created using the IRMA report plug-in v1.3.0.2 (Ion Torrent).

**Table 2.**
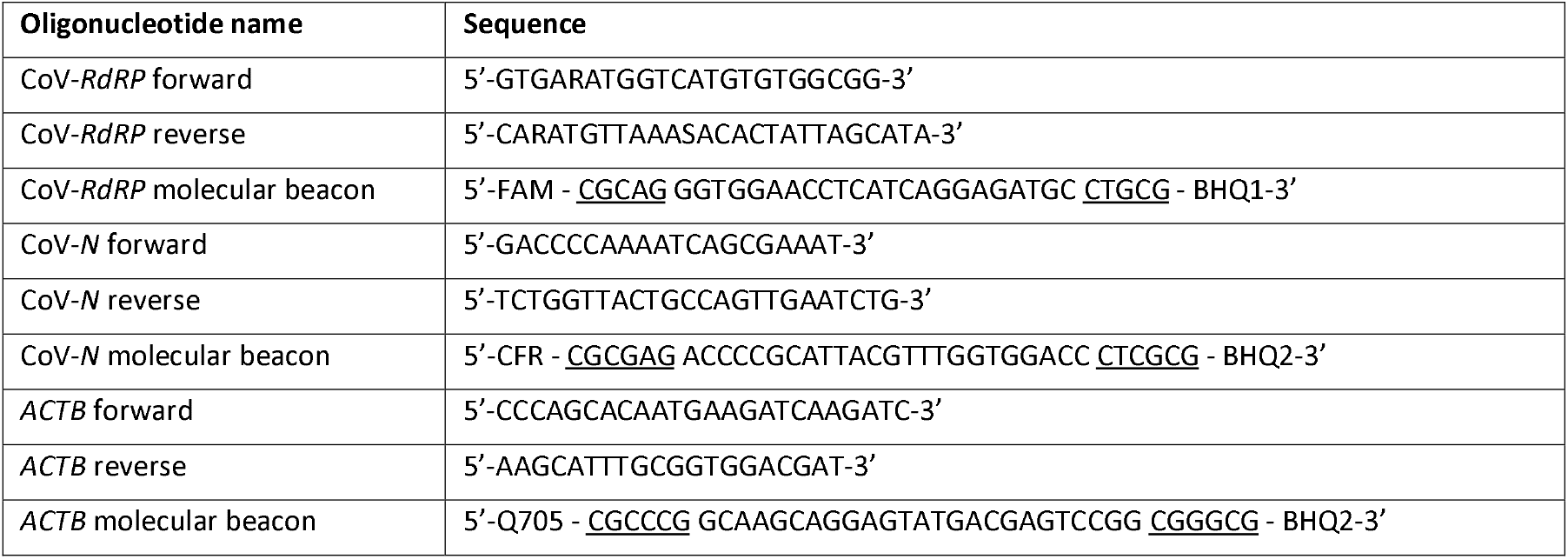
Listing sequences of oligonucleotides used in the SARS-CoV-2 multiplex screening assay presented in Figure S2. FAM = fluorescein, CFR = Cal Fluor Red 610, Q705 = Quasar 705, BHQ = Black Hole Quencher, underlined nucleotides in molecular beacon probes indicate the 5’ and 3’ arm regions of the probe.

Sequence data and corresponding metadata were uploaded to GISAID and are included as supplementary data. Aliquots of purified RNA samples no longer needed for diagnosis were deidentified and transferred to Rutgers’s laboratory for initial testing of the multiplex variant molecular beacon assay.

### RNA Purification from inactivated SARS-CoV-2 Virus

10,000 copies of each inactivated Variant of Concern SARS-CoV-2 virus from ZeptoMetrix (Buffalo, NY) (catalog# NATSARS(COV2)-VP) was treated with proteinase K in a 210 μL reaction (1% SDS, 25 mM EDTA, 10 mM Tris pH8.3 and 20 μg proteinase K (Thermo Fisher, catalog #25530049)) at 45°C for 30min, to decrosslink the protein matrix entrapping the viral RNA. After the addition of 1-2µg of yeast carrier tRNA (Roche, Basel, Switzerland, catalog #10109495001), purification of the RNA was performed using the QIAgen RNeasy Mini kit (catalog #74104).

### Multiplex qRT-PCR with samples

RNA purified patient and inactivated virus samples were tested in a similar manner to how we tested the assay’s sensitivity and specificity. 5 µL of each sample was combined with 15 µL each completed master mix using TaqPath and run as mentioned before.

### qRT-PCR analysis and plotting

Data generated from the CFX96 Touch Real-Time PCR Detection System was analyzed with Bio-Rad’s CFX Maestro Software Version 2.0, which corrects for fluorescence drift, subtracts the baseline, and fits curves to each qRT-PCR signal. Threshold cycles (C_t_) were determined via a manually specified single threshold of our analyzed data, which was clearly above all negative control signals. For the specificity plots, fluorescent signals were normalized by dividing all values by the highest fluorescent value in each channel for the respective tubes.

## Results

### Design of a two-tube, eight mutation, multiplex qRT-PCR assay for SARS-CoV-2 Variants

In our assay, SARS-CoV-2 variants were identified by probing the coding region of the spike protein transcript for the presence of eight different point mutations, as detailed in Figure 1. Since only the mutants are expected to yield a positive signal, we included an additional set of primers and molecular beacons for a conserved region of the viral *N* gene in Tube one as a positive control for the presence of the virus. Since the multiplexing capacity of common thermal cyclers is 4 or 5 colors, we divided the assay into two tubes. The colors of the molecular beacons and the locations of the primers are indicated in Figure 1A. In Tube one, there were three amplicons, and in Tube two, all four targets were present in the same amplicon.

**Figure 1.**
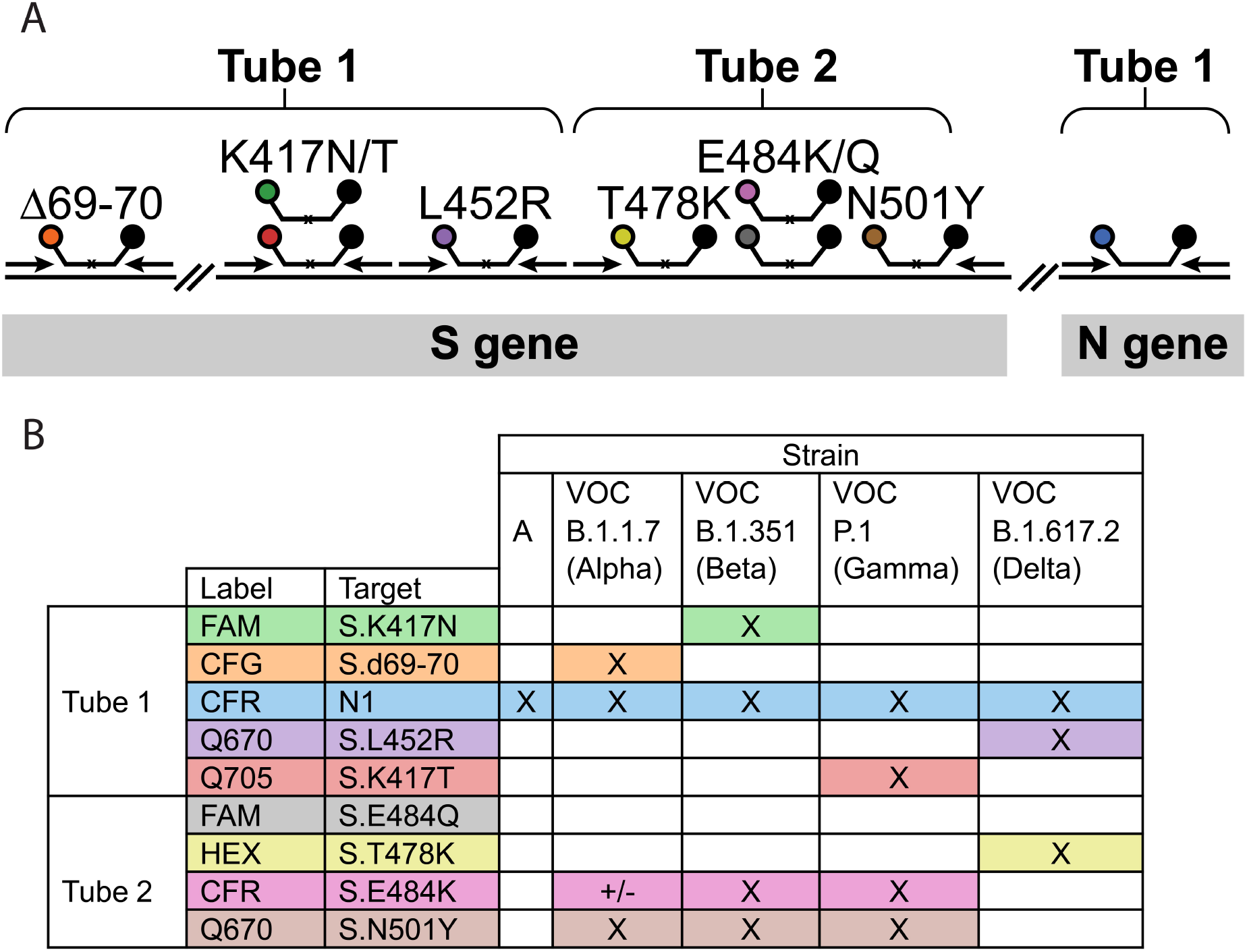
**A**. A schematic representation of the primers and molecular beacons used in the assay. A small black x on the molecular beacons target binding regions indicates that the molecular beacon is single nucleotide discriminant. **B**. Identifying key mutations present in each variant. The mutations detected in the assay are shown in the rows and variants are shown in the columns. Each of the variants of concern can be identified by these combinations of mutations as well as the variants of interest, see supplemental chart. Abbreviations used: CFG = Cal Fluor Gold 540, CFR =Cal Fluor Red 610, Q670 = Quasar 670, Q705 = Quasar 705.

Even though individual isolates of each variant may possess other mutations, we focused on sets of mutations that are functionally the most important. By using *in vitro* binding and epidemiological data of how these mutations affect immune evasion and binding to the ACE2 receptor^2–5,17,18^, we chose to identify the mutations shown in Figure 1: d69-70, K417N/T, L452R, T478K, E484K/Q, and N501Y. We also chose these mutations such that a single mutation in Tube one is sufficient to identify the VOCs, which of course is subject to change as the virus evolves. The mutations targeted in Tube two complete the identification of the VOCs and identify other common mutations, which are useful for identifying other variants.

### The variant molecular beacon assay is specific for the identification of common SARS-CoV-2 variant mutations

Before assembling our multiplex assay, we designed molecular beacons for each targeted mutation, determined their thermal denaturation profiles, and tested them in monoplex PCRs. To determine whether the molecular beacons were allele discriminating at the annealing temperature, we determined the thermal denaturation profiles of the molecular beacons by themselves, and together with mutant or wild-type targets. In these experiments, the targets were synthetic oligonucleotides, and they were used in a molar excess above the molecular beacon concentration. A representative profile is shown in Figure 2, and the profiles of all the molecular beacons are shown in Supplementary Figure 2. Although, the window of discrimination varies somewhat from molecular beacon to molecular beacon, all of our molecular beacons permitted single nucleotide discrimination (see Figure 3A). After qualifying the molecular beacons in this manner, we performed a set of monoplex PCR assays for each mutation, using wild-type targets and mutant targets in alternate reactions.

**Figure 2.**
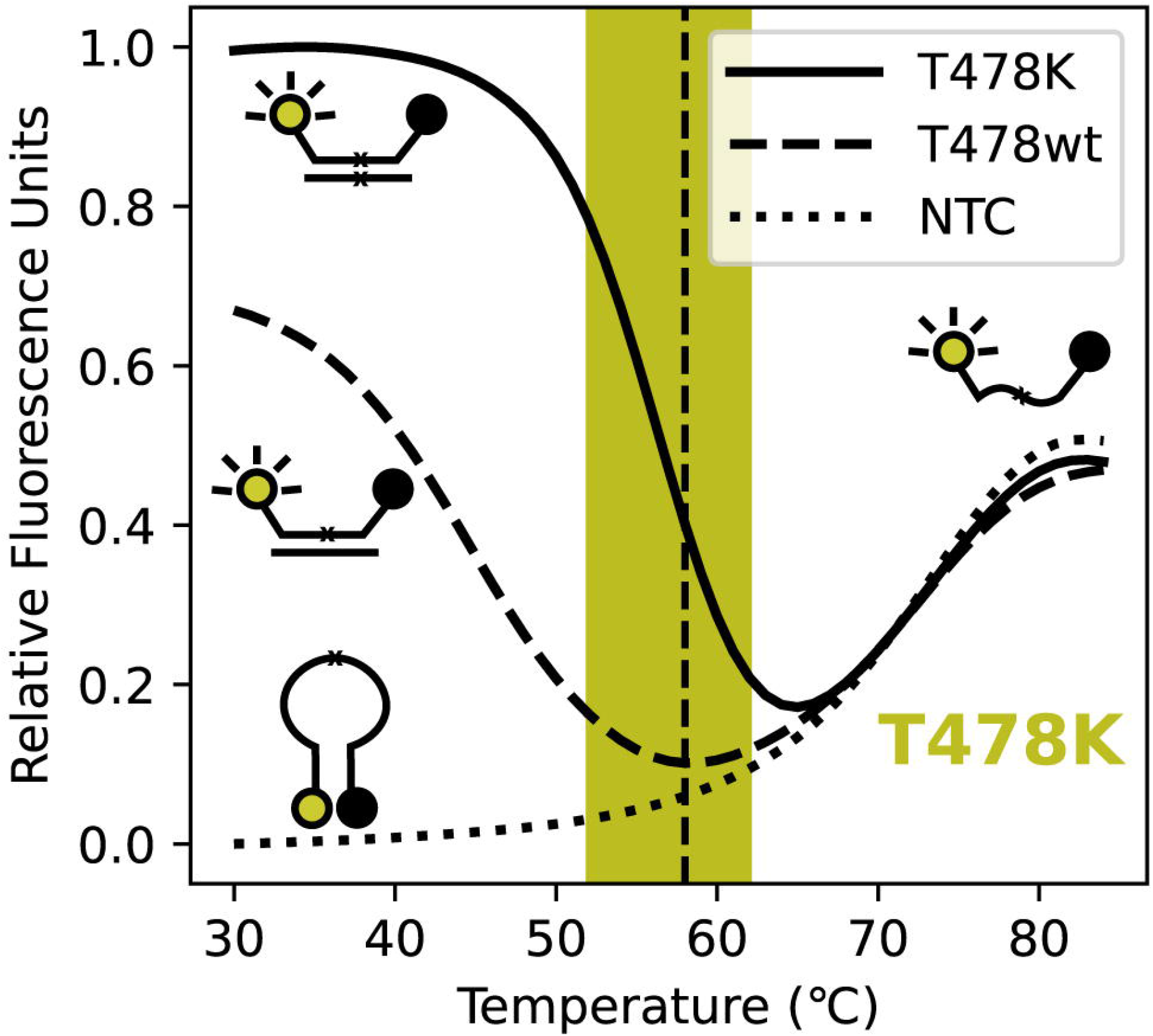
The florescence of the T478K discriminating molecular beacon, as a function of temperature, in the presence and absence of targets. When no target is present, dotted line, the fluorescence increases as temperature rises due to the helical order of the stem giving way to a random-coil configuration, separating the fluorophore from the quencher, and resulting in fluorescence. When the molecular beacon binds to its target its stem dissociates which turns on its fluorescence. Due to the higher binding strength of the perfect probe-target hybrid, solid line, compared to the imperfect probe-target hybrid, dashed line, the imperfect-probe target hybrid dissociates at a lower temperature. The PCR monitoring temperature (58°C) is indicated by the vertical dashed line and the window of good discrimination by the shaded area.

**Figure 3.**
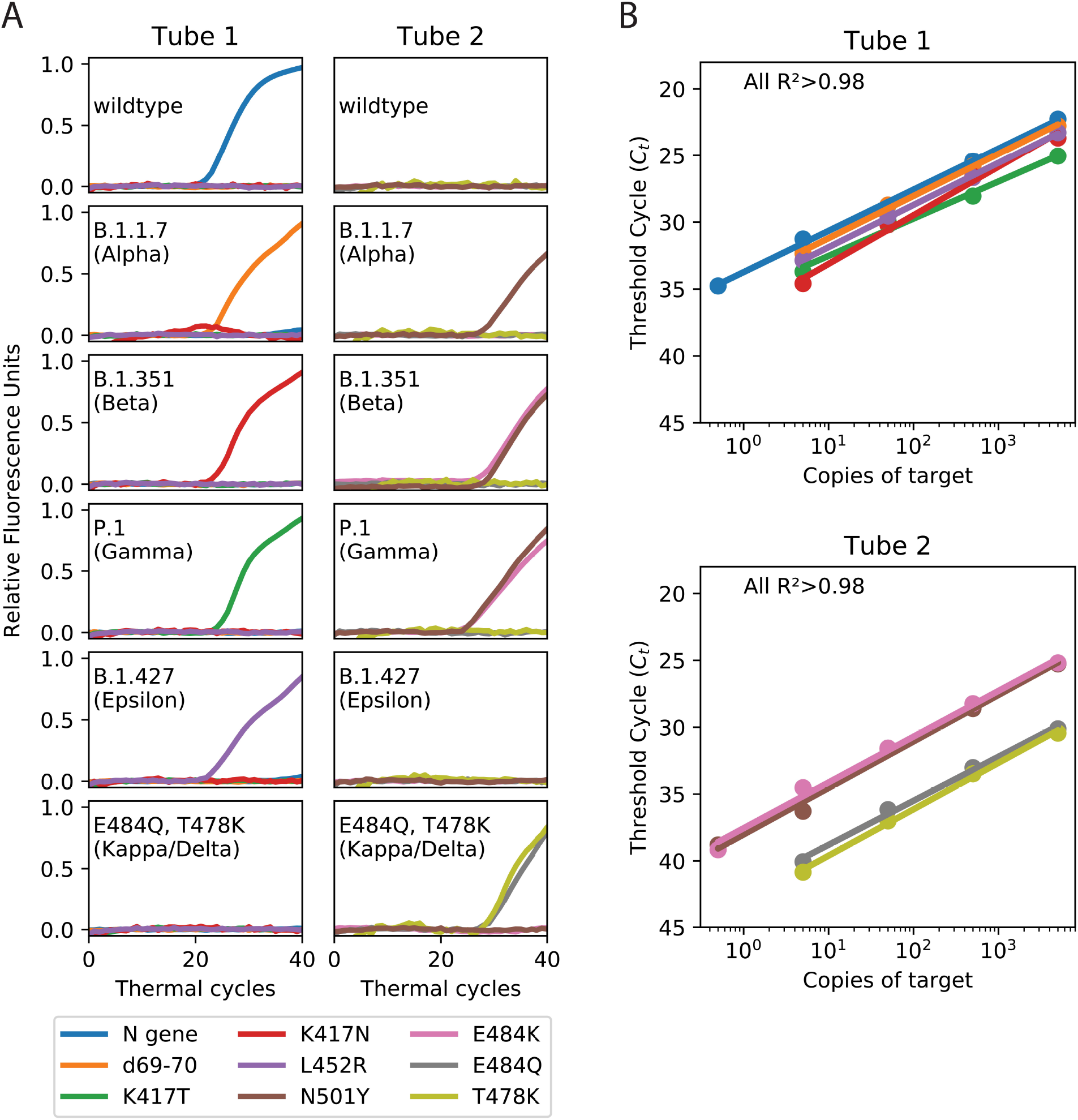
Demonstration of specificity and sensitivity of the variant molecular beacon assay. **A**. Each panel shows the amplification curves of a single multiplex reaction using either the Tube one or Tube two specified primers and probes (columns), and the indicated *in vitro* transcribed RNA targets (rows). The ‘wildtype’ template is RNA of the original strain of SARS-CoV-2’s entire genome. The labeled strains contain the RNA sequences of the S gene from the respective SARS-CoV-2 variants and the sample labeled Kappa/Delta uses an RNA corresponding to (positions 1251-1630nt in the Spike protein coding sequence) and containing the E484Q and T478K mutations, which are from the B.1.617 (Kappa) and B.1.617.2 (Delta) variants. This Kappa/Delta Spike RNA fragment was generated and used due to the lack of commercially available *in vitro*-transcribed RNA’s at the time. **B**. Sensitivity analysis was run by diluting templates containing the specified mutations from 5,000 copies down to 0.5 copies per reaction. The C_t_ values are plotted against the template copy number and the correlation coefficients of the natural log of the copy number to the C_t_ are greater than 0.98 for all amplification products, with around a 3 C_t_ delay between 10-fold dilutions, both as expected.

When all the monoplex reactions indicated successful amplification and allele discrimination, we assembled two multiplex reactions that included all the primers and molecular beacons, as detailed in Figure 1. To demonstrate the specificity of the multiplex assays, six pairs of reactions were carried out, in which each reaction received 5,000 copies of either the wild type, B.1.1.7 (Alpha), B.1.351 (Beta), P.1 (Gamma), B.1.427 (Epsilon), or E484Q and T478K (Kappa/Delta)-containing spike RNA sequences. The targets were *in vitro*-transcribed RNAs corresponding to a portion of the spike protein, or commercially available RNA standards, as detailed in methods and in the legend to Figure 3. The results of these reactions are presented in Figure 3A, where the targets are indicated inside the panels, and the color response of each molecular beacon is indicated by the color of the line. This data shows that when the wild-type SARS-CoV-2 genome is present, only the N gene positive control signal occurs. When testing spike RNAs containing the variant mutations, the correct mutations were identified. For example, the Beta variant (B.1.351) contains not only the uniquely identifying K417N mutation, but also the N501Y and E484K mutation.

### The variant molecular beacon assay can detect at as few as five copies of viral RNA

To demonstrate the sensitivity of the variant molecular beacon assay, PCR assays were initiated with serial dilutions of each target. The results are shown in Figure 3B. Each dilution series tested from 5,000 copies to 0.5 copies. Where a clear signal was present, the threshold cycle was determined by a manually identified single threshold for each channel. Reactions starting with as little as five copies yielded a detectable signal in each case. The correlation coefficients between the logarithm of the copy number and the threshold cycles in each case are each greater than 0.98, showing a very strong exponential relationship between the two. These results also indicate that in Tube one, which contains four pairs of primers, the amplification of multiple different amplicons does not interfere with each other, which would diminish sensitivity.

### Inactivated virus and patient sample strains of SARS-CoV-2 are accurately identified

After demonstrating the specificity and sensitivity of the variant molecular beacon assay with *in vitro*-transcribed RNAs, we performed this assay on RNA purified from inactivated virus obtained from ZeptoMetrix (Buffalo, NY). As can be seen in Figure 4A, our assay identified each of the mutations as listed in Figure 1B for the respective VOC. We next tested a set of patient samples (RNA extracted from mid-nasal swabs) that were SARS-CoV-2 positive, as identified by a multiplex screening assay, which we previously developed (Figure S2). After being identified as positive for SARS-CoV-2 by this assay, the samples were sequenced to identify all mutations present in the strains’ genomes, this sequencing data is available online at https://data.mendeley.com/datasets/v3n2dzt8st/1. Afterwards, the RNA extracted from the samples were then provided to Rutgers in a blinded manner for variant identification.

**Figure 4.**
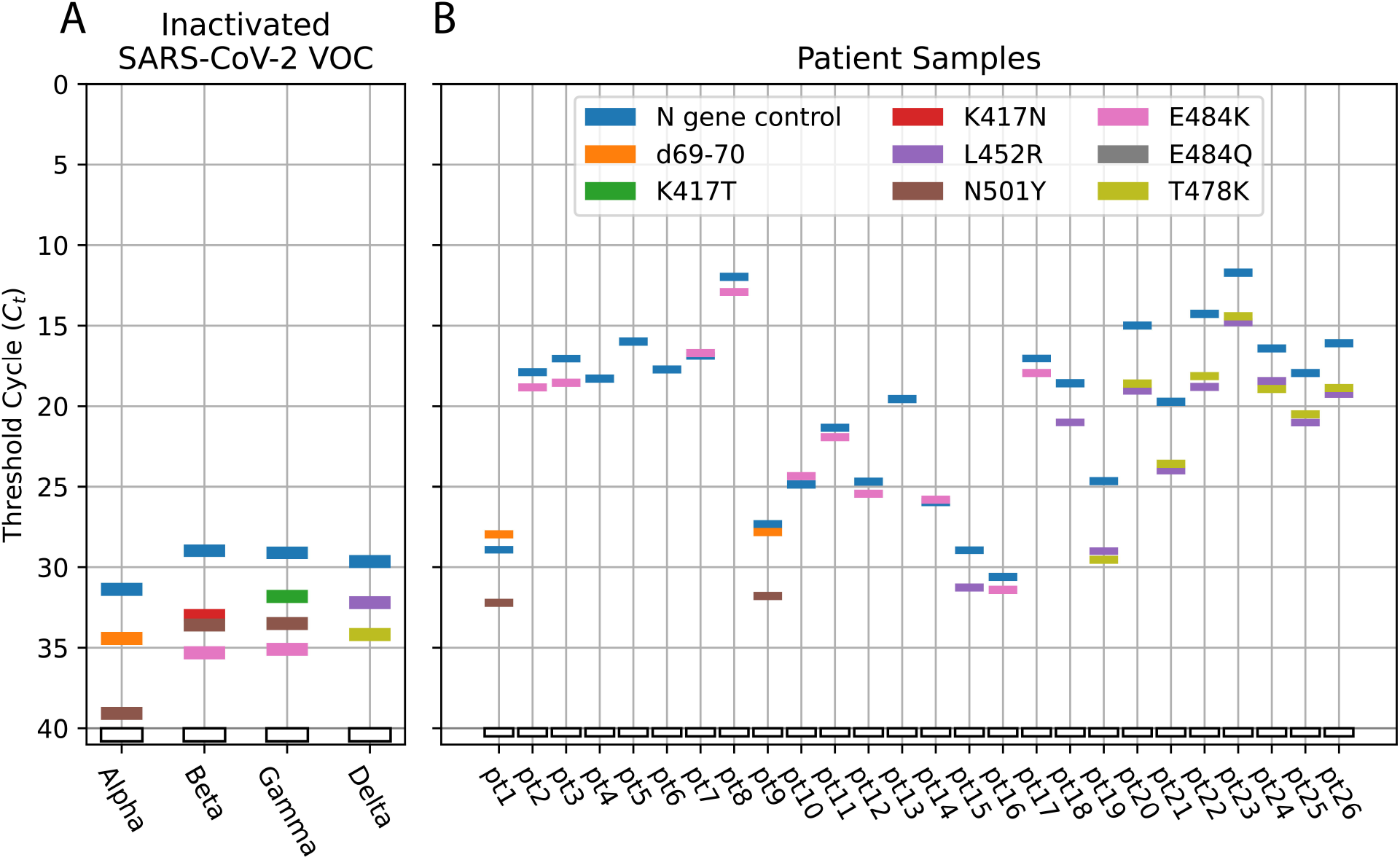
Threshold cycle (C_t_) data for virus and patient samples using the variant molecular beacon assay. **A**. Virus samples. All VOC virus samples tested were accurately identified as shown in Figure 1B. **B**. Patient samples. All samples test positive for the *N* gene control. For the other targets, a C_t_ value less than 40 indicates the presence of the specified mutation, which can be used to identify the variants of the virus. For example, the presence of the T478K and L452R in pt19 indicates that this virus is the Delta variant. All the specified mutations shown in this figure have been sequence confirmed by deep sequencing.

These RNA samples were tested using the two-tube variant molecular beacon assay, as described above. Figure 4B shows the threshold cycles for each molecular beacon target in each sample. All samples tested positive for the *N* gene positive control signal. Two samples were identified as the Alpha variant (B.1.1.7 with d69-70 and N501Y), two samples were identified as the Epsilon variant (B.1.427/429 with only L452R), eight samples were identified as the Delta variant (B.1.617.2 with L452R and T478K), ten samples had the E484K mutation, although not associated with any VOCs, and four samples had none of the targeted mutations and therefore are not any of the currently identified VOCs.

After strain identification was performed with the variant molecular beacon assay, the results were compared with sequencing data, which was in complete agreement. Our results indicate 100% sensitivity and specificity of the assay as tested so far. It is also notable that one of the samples possessed an L452Q mutation, which is similar to the L452R mutation, but the L452R molecular beacon did not show a fluorescent signal due to the L452Q mutation, further confirming the specificity of the results of our single nucleotide discriminating molecular beacons.

### Design and implementation of an Omicron identifying molecular beacon assay

The recent emergence of the omicron variant, which possesses over 30 mutations in the spike protein, necessitates the design of a new identifying single nucleotide discriminating molecular beacon. Since a number of Omicron mutations occur in the target regions of the two-tube assay, it is not feasible to incorporate uniquely Omicron specific molecular beacons into the two-tube assay discussed above in a straightforward manner. Therefore, a new two-color Omicron specific assay was designed by targeting the E484A mutation, which is unique in the Omicron variant (represented in Fig. 5A), along with N gene detection. Previous studies have also shown that this mutation is important for immune evasion^19^. The melt curve for the designed molecular beacon is shown in Figure 5B, which shows high specificity for its intended target at the annealing temperature of the assay of 58°C. Next the sensitivity was tested using *in vitro*-transcribed Omicron spike fragment RNA, similar to what was done previously. The results in Figure 5C show the sensitivity to detect 50 copies of the RNA target. The sequences of probes and primers are listed in Table 3 and the amplicon schematic is shown in Figure 5A. This Omicron assay was tested against the other VOC sequences and the results in Figure 5D show clear identification of the Omicron variant, with no signals from any other target sequences. We then tested thirty-three recent patient samples identifying seventeen of them as Omicron (Figure 5E). These samples were also analyzed using the TaqPath COVID-19 Multiplex Diagnostic Solution assay (ThermoFisher, Waltham, MA) in which Omicron variant fails to produce a S gene specific signal because one of the primers binds in the Omicron variant’s d69-70 mutation. This S gene dropout data was provided to Rutgers in a blinded manner and upon comparison it was found to be in complete agreement, indicating high accuracy of our assay. However, in contrast with TaqPath COVID-19 Multiplex Diagnostic Solution assay our assay yields a positive signal for a unique mutation in the Omicron variant.

**Table 3.**
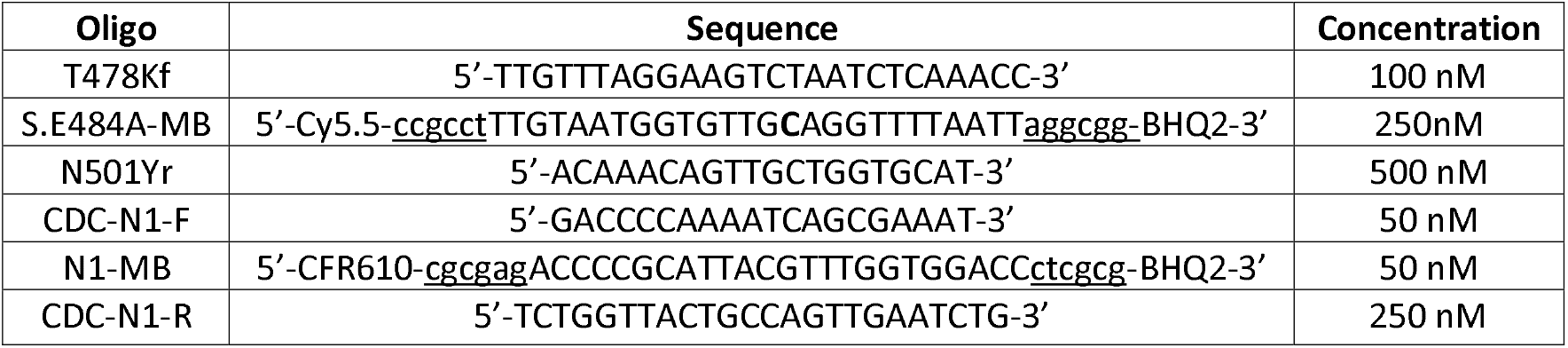
Primers and probes used in Omicron assay. Sequences are shown in the 5’->3’ direction. Upper case text indicates that it is a binding sequence, underlined text forms the hairpins of the molecular beacons, and bold letters indicate the point mutation nucleotides being identified by that molecular beacon. Abbreviations used: CFR = Cal Fluor Red 610, BHQ = Black Hole Quencher

**Figure 5.**
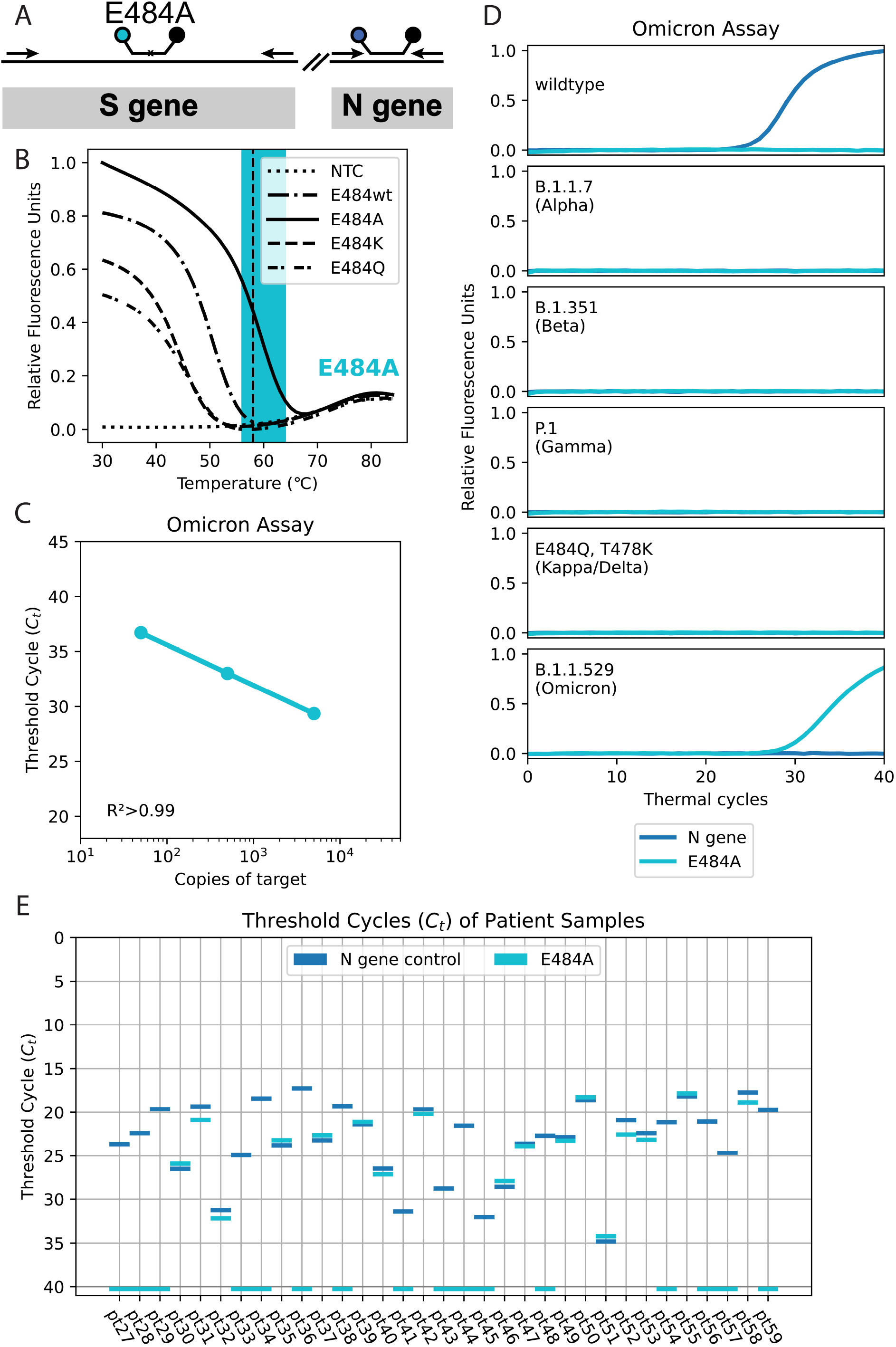
Duplex assay for the Omicron variant. **A**. Schematic showing the locations of primers and molecular beacons. **B**. Thermal denaturation profile of the molecular beacon with various target sequences. **C**. Linearity of detection of *in vitro*-transcribed RNA corresponding to a portion of Omicron S gene. **D**. Specificity of the Omicron assay using synthetic targets (described in Figure 3). **E**. Analysis of 33 recently collected SARS-CoV-2 samples by the duplex omicron assay.

## Discussion

Molecular beacons designed to differentiate point mutations are an ideal tool for genotyping on a global scale due to their ease of design and production. In the case of newer emerging variants, molecular beacons can quickly be synthesized and validated for that variant’s identification, which can help to monitor strains, and improve patient care via epidemiological tracking. Utilizing the outlined strategy, molecular beacons for new targets can be designed and added to the assay in two or three weeks. This assay not only enables the identification of the four variants of concern and many variants of interest, but since it targets many known mutations that confer increased transmission and immune evasion, it will be able to identify new variants that recombine with existing mutations. An example of the assay’s ability to pick up newer variants lies in the Delta variant’s mutations, which include the L452R mutation that is also present in the Epsilon variant as can be seen in Figure 1B.

Due to the specificity of the molecular beacons, other mutations in the target region do not elicit false-positive signals. Evidence of this is found in the lack of signal in one of the samples which contained the L452Q mutation, as identified by sequencing. More evidence of the specificity is in the clean differentiation of the K417T and K417N mutations, as well as the E484Q, E484K, and E484A mutations, where these signals only become positive in the presence of their respective mutations. This specificity enables precise identification, but also comes with the downside that mutations in the binding region of the molecular beacon could result in false-negative signals. Despite this, if other mutations do arise near the uniquely identifiable mutations, such as in the case of the Omicron variant, this can be accounted for by either designing different molecular beacons, or by introducing degenerate nucleotides into the current molecular beacons.

This study is limited by the number of patient samples tested. In the cases where a variant harbors multiple mutations in the target regions of molecular beacons of the two-tube assay, false negatives may result due to the high specificity of molecular beacons. However, in such situations a completely new assay can be easily assembled with equal ease as was demonstrated in the case of Omicron variant in Figure 5.

This assay introduces the sequences of many useful molecular beacons for the identification of point mutations in SARS-CoV-2. These variant molecular beacon assays are most useful as a screening tool for positive SARS-CoV-2 patient samples which can identify all major VOC (see Figure 1B). The primers and molecular beacons can also be combined in varying combinations to test for other pathogens, they can be added to current assays, and they can be used with human RNA controls as were added in the multiplex COVID-19 multiplex screening assay (see Supplementary Figure S2). These assays can then be used to test for the most common variants in a region or to directly test human samples as a single pathogen/COVID-19 variant identification test in general practice.

Here we describe multiplexed molecular beacon assays for the classification of SARS-CoV-2 variants via their identifying functional mutations. We have shown that the assay clearly differentiates between the different SARS-CoV-2 variant sequences, and accurately classifies patient samples that have been identified by deep sequencing technologies and S gene dropout assays. All primer and probe sequences used in this assay are listed in Tables 1 and 3 to facilitate SARS-CoV-2 variant genotyping on a larger scale, and to address the public need for variant tracking.

## Data Availability

All sequencing data is available in the mendeley data link. DOI: 10.17632/v3n2dzt8st.1

https://data.mendeley.com/datasets/v3n2dzt8st/1

## Acknowledgements

This research was supported through a grant from NIH R01 CA227291 and a grant from ResourcePath, LLC. We would like to specifically acknowledge Diana Y. Vargas for her help in designing the SARS-CoV-2 multiplex screening assay.

## Ethics Statement

This study used archived, de-identified samples left over from clinical testing. The study received expedited approval with full waiver of consent, Advarra IRB# Pro00058476.

## Data Sharing

All sequencing data is available online through Mendeley Data at https://data.mendeley.com/datasets/v3n2dzt8st/1.

## Authors’ Contributions

Conceptualization and methodology were performed by RJD, SAEM, and ST. AH, AB, AC, AF provided the patient samples which were tested, and the sequencing results they generated were used to validate the assay used. The original draft was written by RJD, and review and editing were performed by all authors. ST and SAEM supervised RJD in running experiments.

