## Supplementary figure legends for "Multiplex PCR Assays for Identifying All Major SARS-CoV-2 Variants"

Supplemental Figure Legends

**Figure S1**. Melt curves for each molecular beacon with the on and off target oligonucleotides. The windows of discrimination are highlighted in the appropriate color and the annealing temperature for the assay (58°C) is indicated via the dashed vertical line. The on-target sequence oligonucleotide reactions are indicated by the solid lines, the off-target sequence oligonucleotide reactions are indicated by the dashed and dot-dashed lines and the no target reactions are indicated by the dashed lines. The differences in these melting temperatures between on and off target binding are what allows each of these molecular beacons to only become fluorescent in the presence of the target sequence.

**Figure S2.** Molecular beacon-based real-time RT-PCR assay of a serial dilution (100,000 copies to 0 copies) of a SARS-CoV-2 RNA transcript in a background of RNA isolate from 100 HeLa cells. The left plot shows the amplification curves of the SARS-CoV-2 RdRp gene (green curves), the SARS-CoV-2 *N* gene (red curves), and the human *ACTB* gene (blue curves). The right plot shows the inverse linear relationship between the logarithm of the number of SARS-CoV-2 constructs (green dots and line for the *RdRp* gene and red dots and line for the *N* gene) and the number of thermal cycles that are required to generate a visible fluorescent signal. The dotted blue line indicates the threshold cycle of the human *ACTB* gene in each reaction. The plots indicate that as few as 1 copy SARS-CoV-2 RNA in a background of RNA from 100 human cells can be reliable detected and identified.

The *RdRp* and *N* genes were chosen as targets to detect lowly and highly expressed transcripts generated by SARS-CoV-2, as well as genomic copies of the SARS-CoV-2 virus.

Real-time RT-PCR assays were carried out in a 20-µl volume that contained 1 x TaqPath 1-step RT-qPCR Master Mix (A28521, Thermo Fisher Scientiﬁc, Waltham, MA), 100 nM CoV-*RdRP* forward primer, 500 nM CoV-*RdRP* reverse primer, 250 nM CoV-*RdRP* molecular beacon probe, 100 nM CoV-*N* forward primer, 500 nM CoV-*N* reverse primer, 250 nM CoV-*N* molecular beacon probe, 100 nM *ACTB* forward primer, 500 nM *ACTB* reverse primer, and 250 nM *ACTB* molecular beacon probe. Each reaction was initiated with 2 to 5 µl RNA template. The PCR assays were performed in 200-µl white polypropylene PCR tubes (USA Scientiﬁc, Ocala, FL) in a CFX96 Touch real-time PCR detection system (Bio-Rad Laboratories, Hercules, CA). The thermal cycler was programed to incubate the reaction mixtures for 10 min at 53°C to generate cDNA, followed by 2 min at 95°C to activate the DNA polymerase and by 45 thermal cycles that consisted of DNA denaturation at 95°C for 15 s and primer annealing and elongation at 58°C for 60 s. Molecular beacon ﬂuorescence intensity was monitored during the 58°C annealing and chain elongation stage of each thermal cycle.
