## Supplementary figures and images for "Multiplex PCR Assays for Identifying All Major SARS-CoV-2 Variants"

### Supplemental Figure S1

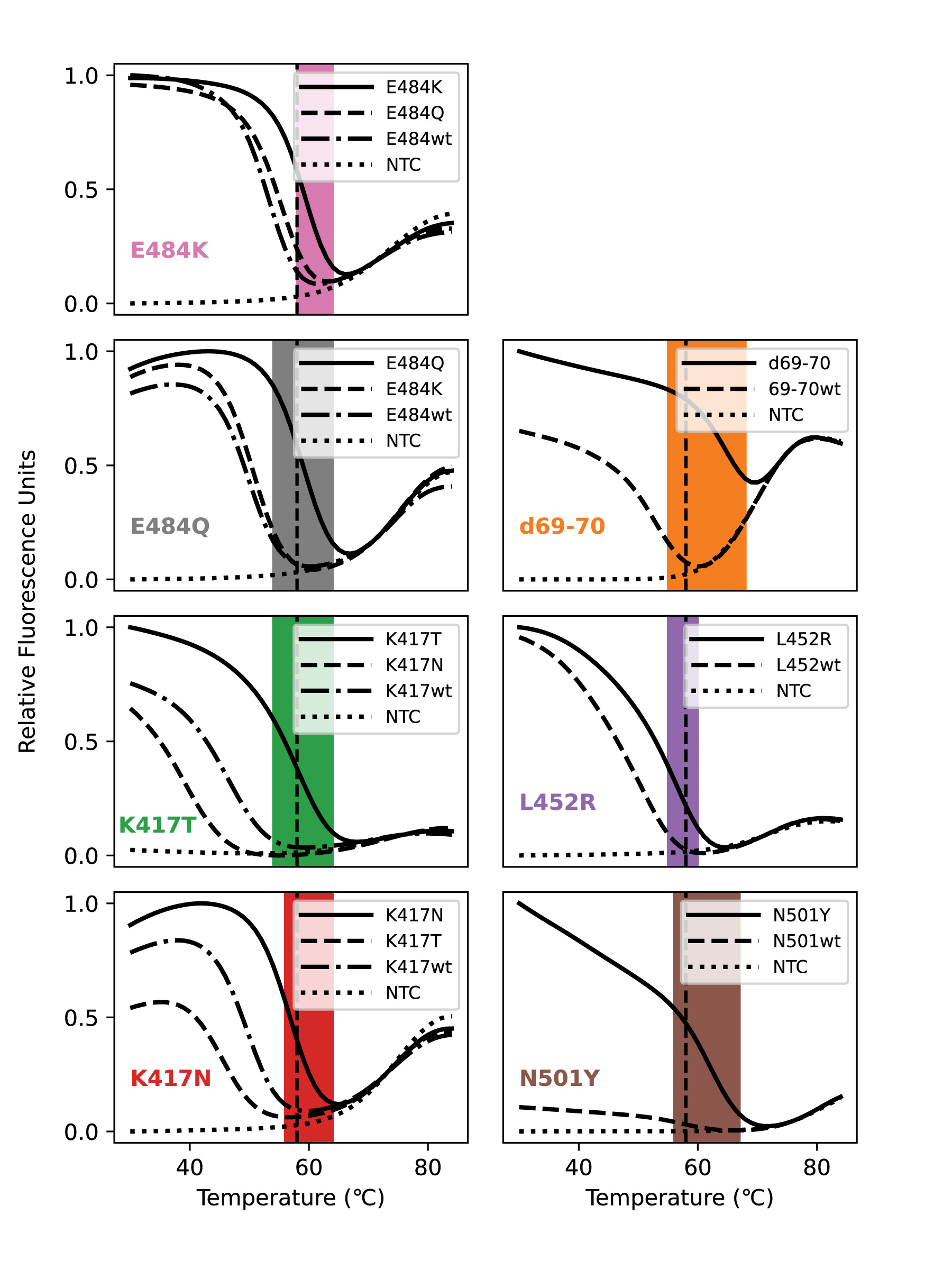

### Supplemental Figure S2

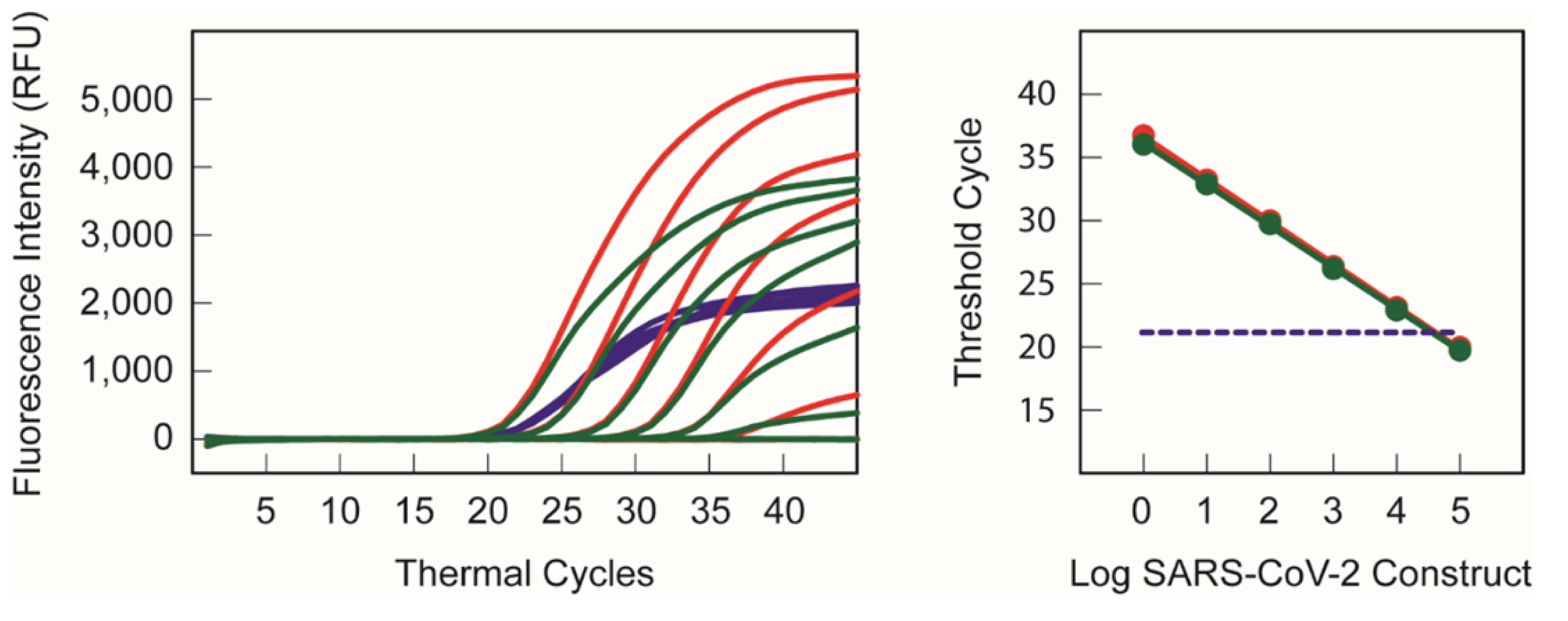
